# How to Flatten the post-lockdown epidemic trajectory

**DOI:** 10.1101/2020.05.06.20093104

**Authors:** Roschen Sasikumar, Ajit Haridas

## Abstract

Populations are locked down during an epidemic to slow down the rate of infection so that epidemic trajectory is "flattened". This helps to keep cases at a manageable level. Given the enormous economic damage and misery caused by a lockdown, it is imperative to keep the lockdown period limited. A lockdown is useful only if it can be ensured that after the lockdown is lifted, the epidemic trajectory does not rise sharply again. We present here the results from a mathematical model of the epidemic which examines how the timing, strength and duration of the lockdown affects the post-lockdown epidemic trajectory. Our results show the following:

1. A early lockdown (imposed when less than 1% of the population has been infected), of any reasonable duration, cannot prevent the return of the epidemic when the lockdown is lifted. The curve starts climbing soon after lifting the lockdown and reaches a peak of the same height as the no-lockdown curve
2. The post-lockdown trajectory can be flattened only if the lockdown is imposed after about 10% of the population has recovered after infection.
3. The slope of the post-lockdown epidemic curve depends on the level of immunity built up in the population before and during the lockdown period. Application of lockdown around the inflexion point of the epidemic curve (the point of maximum slope of the curve) ensures that the post-lockdown curve is also flattened.

## Introduction

Most countries have a strategy of lockdown to manage the SARS-CoV2 pandemic. Since lockdowns come with enormous social and economic costs, it is essential to limit its duration and to ensure that the epidemic peak does not reappear after it is lifted. If the infection reappears after lockdown at the same levels as without the lockdown, the purpose of the lockdown is defeated. Since these mitigation strategies have no precedents to draw upon, the future scenarios must be evaluated through mathematical models.

## The Model

In this study we use a compartment model of the epidemic with 6 compartments – **S**usceptible, **I**nfected (Asymptomatic), **A**iling {symptomatic}, **C**ritical, **R**ecovered, **D**ead. The equations governing the rate of change of population in these compartments are given in APPENDIX 1

We use this model to investigate how the timing of the lockdown affects the post-lockdown infection curve.

Lockdown is represented in the model as a reduction in contacts and thereby a reduction in infection rates. If the no-lockdown infection rate is α0, lockdown is represented by using an infection rate of f α0 where f is a factor between 1 and zero that represents the extent to which the lockdown has reduced normal contacts.

## Results

### Early Lockdown – (Lockdown imposed when less than 1% of the population is infected)

In Figure 1 the blue curve shows the fraction of Infected individuals in the population starting from Day 1 of the epidemic without lockdown and the epidemic is allowed to run its natural course. From Day 1 to Day 70 the fraction infected rises only to 1% of the population but from Day 70 the fraction increases rapidly to a peak of nearly 22.5% of the population in 30 days and then falls to 1% in another 40 days.

The orange curve shows the epidemic curve with mild lockdown (f=0.8, the contacts are reduced to 80% of the normal value) for 20 days starting from Day 20. The green curve shows the epidemic curve with strong lockdown (f=0.2, contacts reduced to 20% of the normal value) imposed during the same period. It can be seen that the shape of the curves with lockdown are identical to the no-lockdown curve except for a delay in the occurrence of the peak. The same result is obtained whether the lockdown is for 20,40,60,80 or 100 days

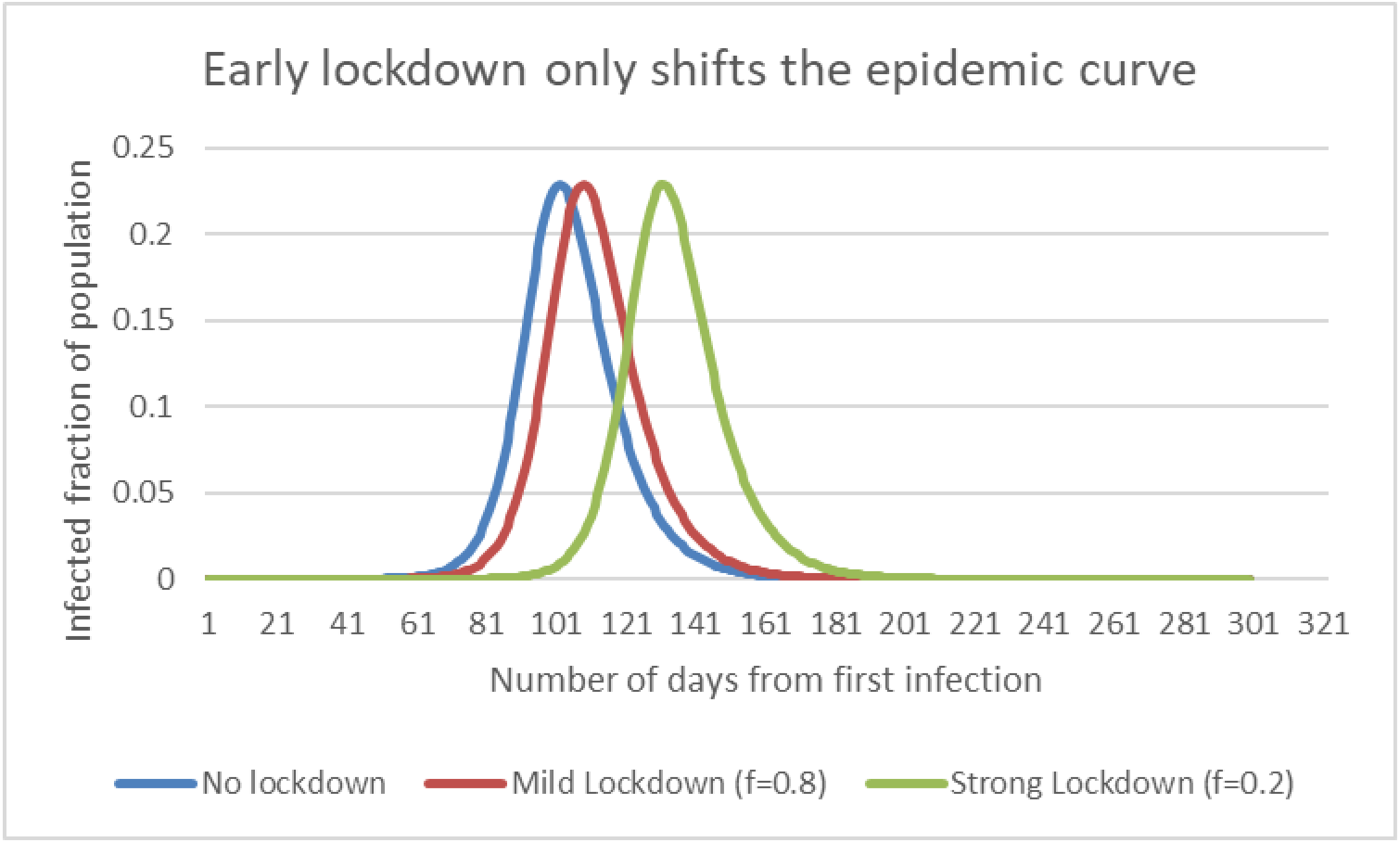

### Later lockdown (after 1% has been infected)

Figures 2 a-c show how the shape of the post lockdown curve depends on the day on which the lockdown is imposed. All the lockdowns are for 20 days starting at different points of the epidemic trajectory. The blue curve is the no-lockdown curve and the red curve is the curve obtained with a lockdown of 20 days.

**Figure 2a.**
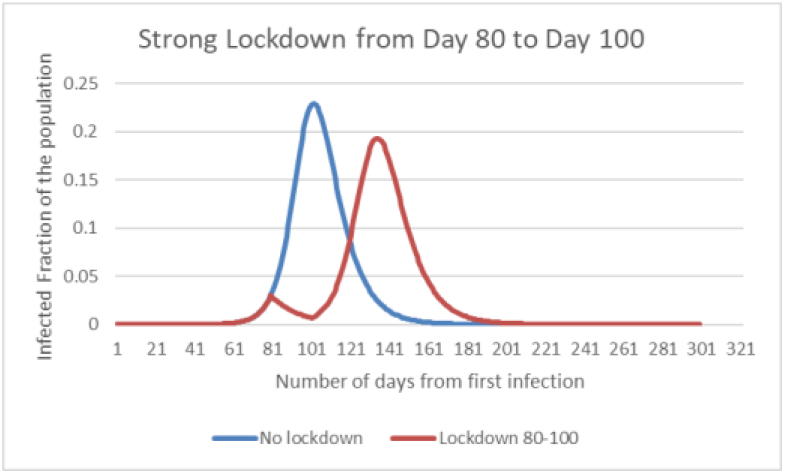

**Figure 2b.**
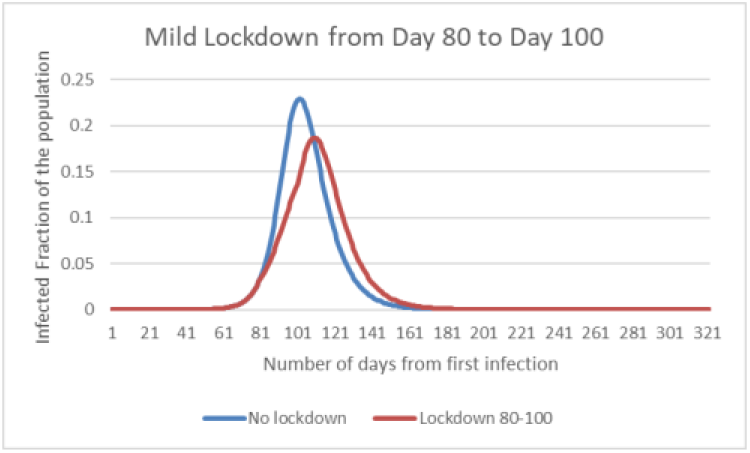

**Figure 2c.**
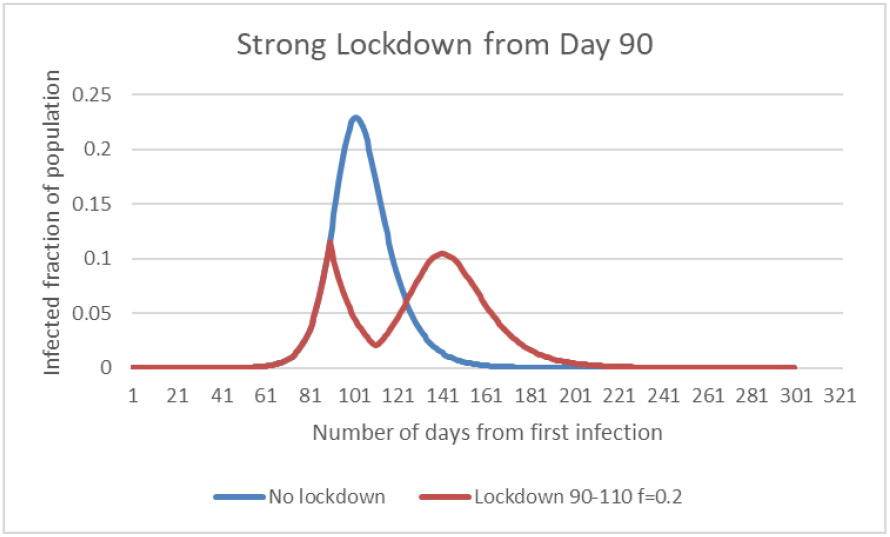

**Figure 2d.**
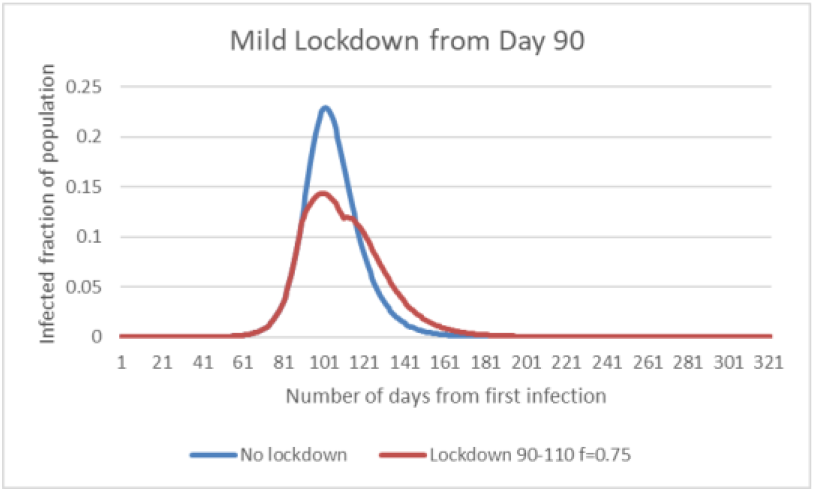

**Figure 2e.**
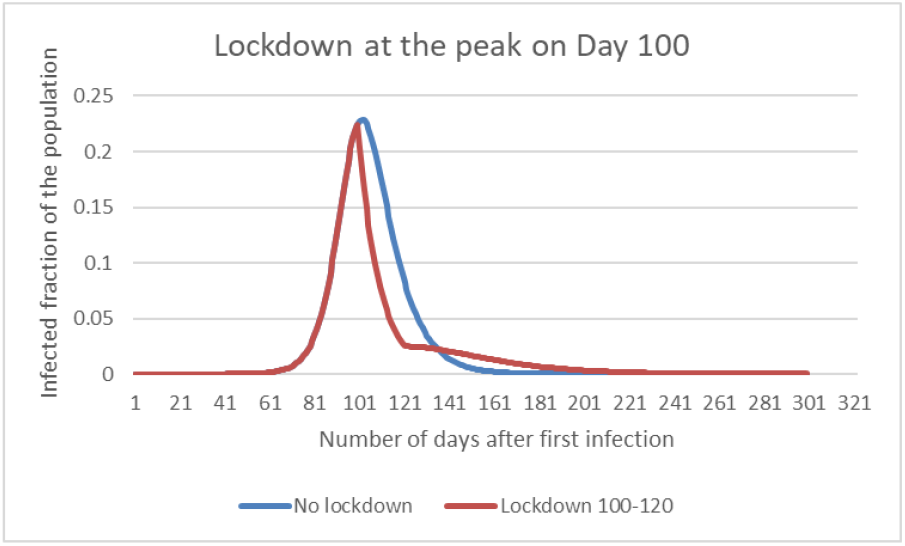

Strong locking down imposed on Day 80 (Fig.2a-b) reduces the peak from 22.5% to 19.3%. Mild lockdown results in a slightly lower post-lockdown peak of 18.6%

Locking down on Day 90 (Fig.2c-d) ensures that peak always stays below 10% in the case of strong lockdown (f=0.2) and below 15% when the lockdown is mild (f=.75). This is maintained even after lockdown is lifted thereby achieving continued flattening of the curve

On locking down on Day 100 (Fig.2e) the peak of the curve is 22.5% and the curve comes down rapidly and the post lockdown curve is flat and low. Locking down after the peak is passed makes very little difference to the curve.

More than the duration of the lockdown, it is the point of imposition of the lockdown that effectively flattens the post-lockdown curve. If lockdown starts from Day 90, 12 days is almost as effective in flattening the post-lockdown curve as a 20 day lockdown (Figure 3).

**Figure 3.**
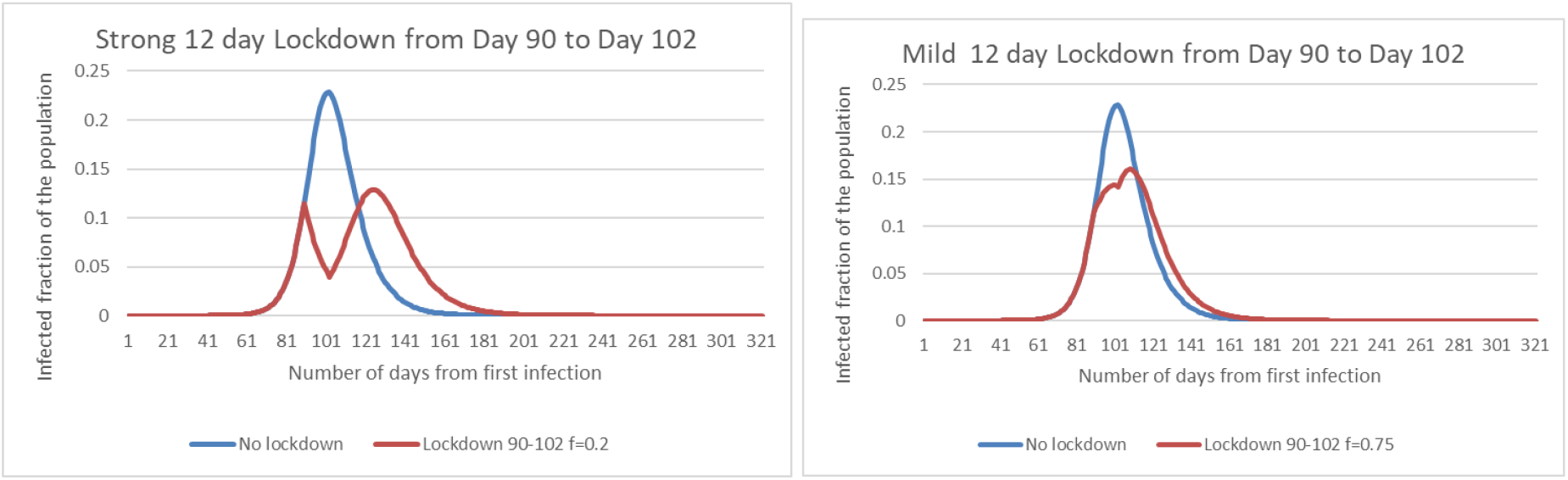

## Discussion

The results show that early lockdowns are totally ineffective for flattening the curve. When the lockdown is lifted the original trajectory comes back with the same intensity. This is true irrespective of how long the duration of the lockdown is.

However when the lockdown is imposed during a narrow window of time points on the trajectory, the post lockdown curve also gets flattened.

The reason for these results can be understood if we observe that the infection rate comes down naturally as more and more of the population recover from the infection and become immune.

Figure 4 shows the variation of infection rate (*dI / dt*) with the fraction of Recovered (immune) in the population. We can see that, in the natural course of the epidemic, it is the developing immune fraction that makes the infection rate slow down. It can be seen from the figure that as the immune fraction becomes greater than about 15%, the infection rate starts decreasing. This is the inflexion point of the infection curve where its slope (*dI/dt*) is maximum. So the infection rate gets lowered naturally when the fraction of immune population corresponds to the inflexion point of the infection curve, in the present case, this happens when the immune fraction is around 15%.

**Figure 4.**
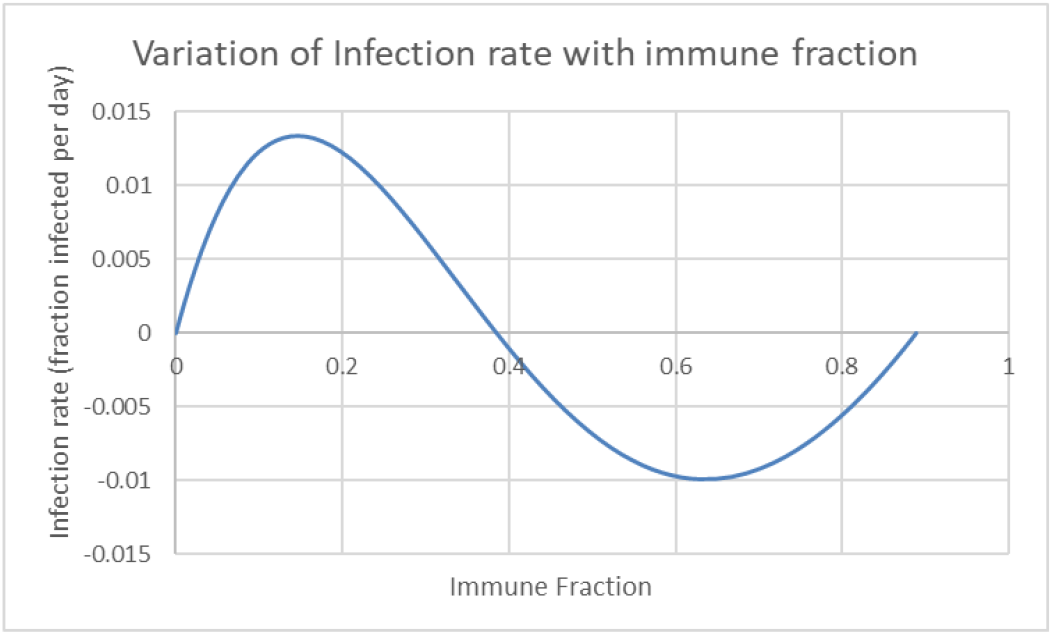

When we slow down infection rate by locking down the population, at a time when the infection rate is very low and hardly any immune fraction has been built up, there will be nothing to slow down the infection rate once the external controls are removed. Therefore the infection resumes at the pre-lockdown rate and no additional flattening is obtained.

On the other hand, if lockdown is imposed close to the maximum infection rate (inflexion point), the already built up immune fraction, along with whatever is additionally built up during the lockdown period, would naturally slow down the infection even after lifting the lockdown and resuming contacts.

In Figure 5 we show the curves for the infected and the Recovered in the case when lockdown was imposed from Day 90 to Day 110 (as in Figure 2 b). In this case the fraction of the Recovered in the population has already reached about 10% when the lockdown is imposed. The immune fraction (the red curve) continues to increase during the lockdown period, albeit more slowly because the infection rate is lowered by the lockdown (the blue curve). So when the lockdown is lifted on Day 110, an immune fraction of over 20% has already been built up in the population. This fraction is large enough to retard the infection rate even after the lockdown is lifted. So the post lockdown curve is also flattened.

**Figure 5.**
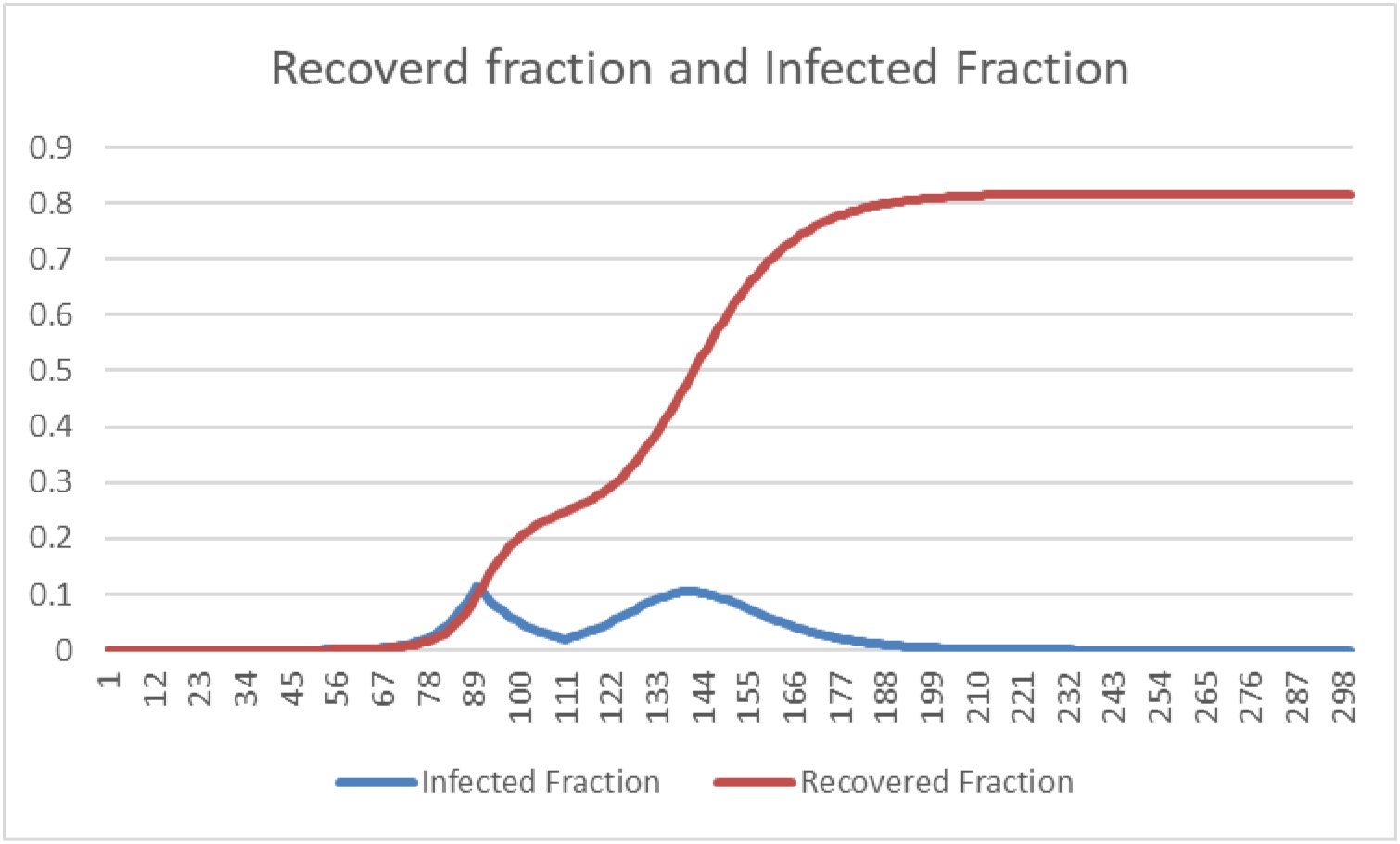

## Conclusion

Lockdowns are effective in maintaining the flattening of the curve after the lockdown period only if the lockdown is imposed after building up sufficient fraction of immune population. This fraction corresponds to the point of inflexion of the epidemic curve where the rate of infection reaches a maximum. Whether it is practical to wait till the point of inflexion to impose lockdown, is a valid question that has to be answered with many other considerations. But any practical solution must work around the fact that, the earlier the lockdown is imposed the higher will be the post-lockdown resurgence.

## Data Availability

No data is referred in the article

## APPENDIX 1

### Model Details

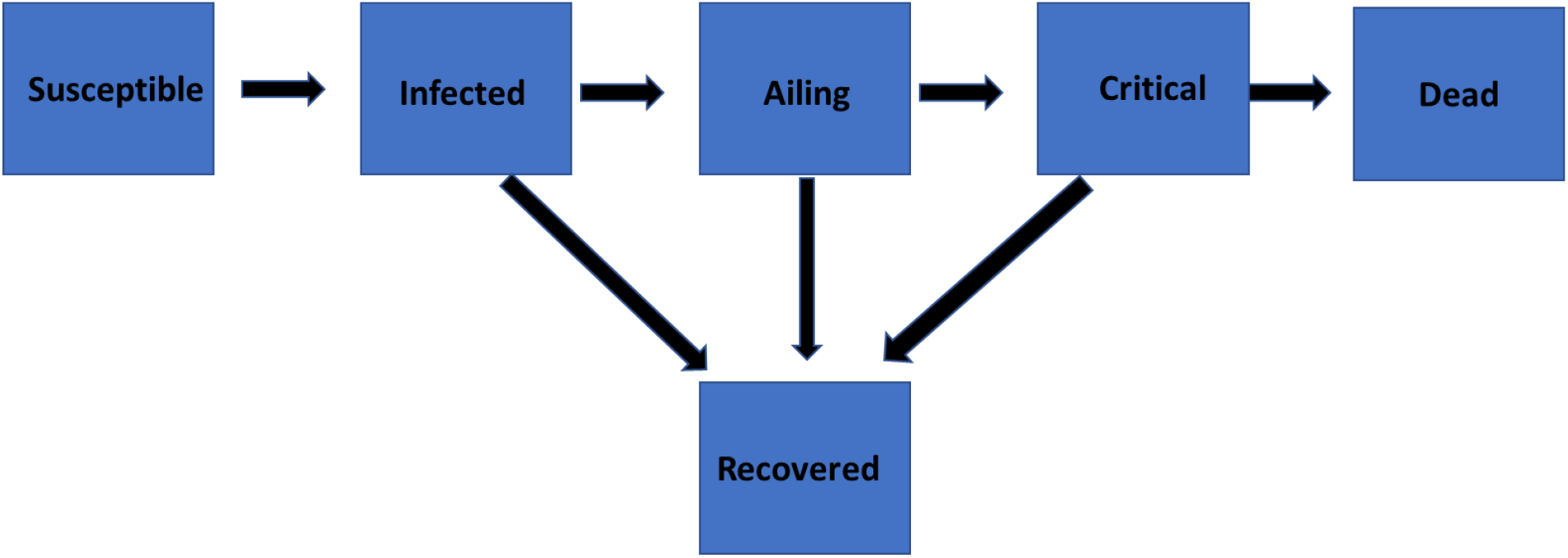

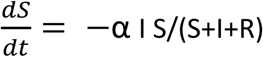

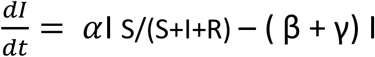

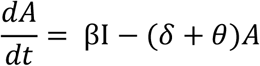

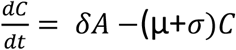

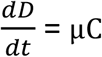

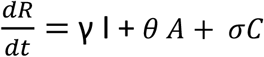

Where:

S = Susceptible population I = Infected population A = Ailing population C = Critically sick population D = Dead population R = Recovered immune population α, β, γ, δ, θ, μ, σ are the rates of transfer between the compartments

**Transfer rate values used in the simulations are given below:**

S-> I rate - α = .3018 I-> A rate - β = .0005 A-> C rate - δ = .0112 C-> D rate - μ = 0.089 I-> R rate - γ =0.12475 S—> R rate − θ = 0.118 C → R rate − σ = .0133

